# Accounting for Human Movement to Improve Exposure-Health Models

**DOI:** 10.64898/2026.06.15.26355663

**Authors:** Hadiqa Tahir, Simon Smart, Samuel Cai, André Ng, Joshua Vande Hey, Tim CD Lucas

## Abstract

**Background:** Current exposure-health models rely on averaged, residential-based environmental exposures, failing to account for human movement. This aggregation can lead to exposure misclassification and biased exposure–response estimates, potentially distorting our understanding of the true health effects of environmental conditions. We developed exposure disaggregation regression models that explicitly account for human movement when linking environmental exposures to health outcomes.

**Methods:** By weighting pixel-level exposures according to distance from home as a simple proxy for human movement, our model linked disaggregated environmental exposures to individual-level health outcomes. Weights were either fixed *a priori* or derived from a latent distance-decay power parameter learned from the data. We additionally evaluated model performance under a nonlinear exposure–response relationship. Model performance was assessed across multiple sample sizes (N = 1,114; 50,000; and 100,000). A simulation study examined parameter recovery using bias, empirical standard error (EmpSE), and credible interval coverage. As a case study, Demographic and Health Surveys (DHS) data from Albania were used to link acute respiratory infection (ARI) outcomes among children under five to pixel-level NDVI within a 3 km buffer around DHS cluster centroids, and the proposed models were applied to these data.

**Results:** Across all models (fixed-weight, learned-weight, and restricted cubic spline models), parameter recovery improved with increasing sample size. At N = 1,114, estimates were biased and imprecise, with incorrect effect direction for exposure-response parameters (e.g., learned-weight *β*_1_ bias = −0.79; EmpSE = 2.61; coverage = 0.88). In contrast, the models accurately recovered parameters at larger sample sizes, including the latent distance-decay parameter (bias = −0.02; EmpSE = 0.15; coverage = 0.95 at N = 100,000), demonstrating their ability to reliably learn movement-based exposure weights when sufficient data were available.

**Conclusion:** Instead of relying on arbitrarily-sized buffers, this statistical framework provides a novel method for studying environmental exposure-health relationships whilst accounting for human movement. With sufficiently large sample sizes, it can accurately estimate the influence of disaggregated environmental exposures on individual-level health and help address exposure misclassification arising from residential-only metrics. This methodological framework remains scalable, interpretable, and adaptable to other exposures and outcomes, offering a foundation for future work that integrates richer mobility-informed exposure-health research.

## 1 Introduction

Understanding the effect of environmental exposures—such as air pollution or green-ness—on respiratory health requires accounting for human movement; where people actually spend their time, not just where they live. Despite this, many epidemiological studies still rely on environmental exposures averaged over a buffer centred on fixed locations (e.g. residence, school, or workplace) (Browning and Lee 2017; Fernandes et al. 2023; Hartley et al. 2020), overlooking how human mobility, land use, accessibility, and built-environmental heterogeneity shape environmental exposure (Oliver et al. 2007) and, ultimately, health. Thus, failing to account for the dynamic environments individuals routinely move through introduces exposure misclassification and potentially biased health findings (Kwan 2018; Kwan 2012).

In exposure modelling, studies comparing dynamic, movement-based exposure with residential-only estimates have found that accounting for human mobility often produces higher or differently distributed exposure levels (Kim and Kwan 2019; Lu 2021; Ma et al. 2020). Smith et al. (2016) developed the London Hybrid Exposure Model, which incorporates time–activity data across indoor, outdoor, and transport environments. They found that accounting for people’s daily movements produced lower average exposure estimates than residential-only measures, largely because individuals spend around 95% of their time indoors where outdoor pollution is reduced, and because transport modes influence exposure patterns. Similarly, Tayarani and Rowangould (2020) computed dynamic PM_2.5_ exposure by tracking each person’s time in different traffic analysis zones and along road links, weighting pollution exposure by time spent in each location and during travel. Dynamic exposure was much higher than home-based exposure—by about 51%—and the degree of exposure misclassification varied across urban, suburban, and rural areas. In another example, Zheng et al. (2024) found that the interaction between travel distance, time outdoors, and irregular movement patterns weakened the link between static and dynamic greenspace exposure, highlighting the importance of accounting for mobility.

Interestingly, although these studies have demonstrated how daily movement can over- or underestimate exposure, only a few have investigated the implications of these estimates for health risk assessment. Emerging studies across several fields suggest that incorporating mobility might improve the representation of environmental exposures relevant to health outcomes. For example, incorporating high-resolution intraday human mobility into *Aedes*–human exposure models improved predictions of dengue risk by capturing when and where people encountered mosquito vectors (Knoblauch et al. 2025); dynamic air-pollution exposure estimates derived from GPS-tracked mobility have been shown to yield stronger associations with biomarkers of metabolic disorders than traditional residence-based exposure measures (Letellier et al. 2022); and ignoring mobility in studying the spatial heterogeneity of thermal conditions may misclassify exposure and bias temperature-related health risk assessment in outdoor workers (Sugg et al. 2019). However, existing studies remain limited in scope, scale, and generalisability. For instance, Letellier et al. (2022) relied on a relatively small cohort (N = 552 participants, 42% Hispanic) and short observation period of two weeks, while Knoblauch et al. (2025) focused on a specific vector-borne disease context rather than broader environmental health outcomes.

Despite the rationale for incorporating human mobility into exposure–health research, this remains challenging for several reasons. First, health (e.g. asthma, COPD), environmental (e.g. NDVI, PM_2.5_) and mobility data (e.g. GPS traces) are typically housed in separate administrative systems and are governed by strict ethical safeguards to protect individuals’ identities. As a result, such data have been difficult to access and link to health outcomes, limiting the extent to which mobility could be incorporated into exposure–health research. In recent years, however, a growing body of work has begun developing open-access anonymised approaches for sharing georeferenced mobility data—for instance, large-scale de-identified mobility records for urban research in England (Zhong et al. 2025), US-wide collections of anonymised activity spaces derived from geotagged social media (Poorthuis et al. 2024), activity space maps for quantifying time spent at risk generated from mobile phone user app location history (Citron et al. 2021), and Safegraph Patterns mobility data (Li et al. 2024). These data mark an important step toward mobility-informed environmental and health research.

Second, integrating health, mobility, and environmental data pose methodological challenges. Mobility and environmental exposures are measured at fine spatial resolutions, whereas health data are recorded at the individual level. Linking these requires statistical frameworks capable of addressing spatial misalignment and the multiscale structure of the data (Banerjee et al. 2003). In practice, however, these challenges have often been managed by aggregating to coarser spatial or temporal scales, so they align with health data. While this approach facilitates analysis, it obscures how disaggregated ‘spikes’ in pollutant levels or abrupt environmental conditions may disproportionately influence physiological responses (Chandia-Poblete et al. 2022).

Finally, analyses that integrate high-resolution mobility, environment, and health data are computationally demanding, particularly when applied to large populations or long time periods. Together, these barriers help explain why relatively few studies have examined the health implications of mobility-informed exposure assessment. This area of study remains an important gap in the literature, especially for respiratory health.

Respiratory conditions, such as asthma, COPD, and acute respiratory infection (ARI), are sensitive to environmental conditions that vary over short spatial and temporal scales (Idrose et al. 2022; Su et al. 2024). Greenness, for example, is a complex environmental exposure as it involves both protective and harmful pathways in respiratory health research (Johannessen et al. 2023). It may improve air quality by filtering pollutants, foster microbial diversity, and promote physical activity (Nieuwenhuijsen et al. 2014; Chen et al. 2017), yet it can also generate allergenic pollen or trap pollutants depending on vegetation type, diversity, and seasonality (Kim et al. 2025), potentially exacerbating respiratory symptoms in some individuals. How individuals move through, or spend time within, these heterogeneous green environments may further shape their exposure in ways that are either beneficial or harmful.

Here, we use greenness and acute respiratory infection as a case study to demonstrate how simple human movement models can be incorporated into exposure estimation. In this study, our goal is not to estimate the real-world association, but rather to use this setting as a test for our modelling approach. We develop and test a statistical method for incorporating human movement into exposure-health models. While the method is broadly applicable to different exposure–outcome combinations, we demonstrate its implementation using disaggregated NDVI data, weighted according to a simple model of human movement, in a simulated ARI context. The aim of this study was to assess model performance under varying sample sizes and structural complexities to understand its capacity to recover parameters.

## 2 Methods

This study used multiple data sources—health data on ARI, geographic data, environmental exposure data on NDVI—along with a simple movement model constructed to approximate how individuals interact with their immediate environment.

### 2.1 Health and geographic location data

The children’s recode of the Albania Demographic and Health Survey (ADHS), collected between 11 September 2017 and 20 February 2018 (Institute of Statistics et al. 2018), was obtained and restricted to urban households as GPS displacement error is smaller in urban clusters. Geographic data for ADHS cluster centroids were provided as georeferenced coordinates that were randomly displaced by up to 2 km in urban areas to protect participant confidentiality. After excluding children with missing health information or those reported deceased, the final analytic sample comprised 1,114 urban children.

Acute respiratory infection (ARI) was defined following ADHS guidelines. Children under five were classified as having ARI if their mothers perceived fast or difficult breathing due to a chest-related problem in the two weeks preceding the survey.

### 2.2 Environmental data

Normalized Difference Vegetation Index (NDVI) data were acquired from Google Earth Engine at a 100m x 100m spatial resolution. To minimise the influence of cloud cover, particularly in Sentinel-2 imagery, an annual median NDVI composite was generated for 2018 due to limited temporal information on DHS data. NDVI was used as the sole environmental metric given the methodological case study focus of this work. For each DHS cluster centroid, pixel-level NDVI values within a 3,000-m buffer were extracted using the *sf* and *terra* packages in R. This buffer size was selected to ensure coverage of all households contributing to each cluster. Because DHS households within a cluster share a centroid location, children from the same cluster received identical NDVI values.

### 2.3 Modelling Human Movement

To incorporate mobility, we assumed a simple distance-decay movement model, in which the probability of a child visiting a given pixel decreases with distance from home. Pixel-level NDVI values were therefore weighted using an inverse distance function, giving greater weight to pixels closer to the cluster centroid (see Figure 1). This reflects the idea that environmental hazards in the immediate residential surroundings are likely to influence health more strongly than areas farther away. These weights can be altered to consider the time spent, probability of visiting particular pixels, or a combination of expectation of time visiting.

**Figure 1:**
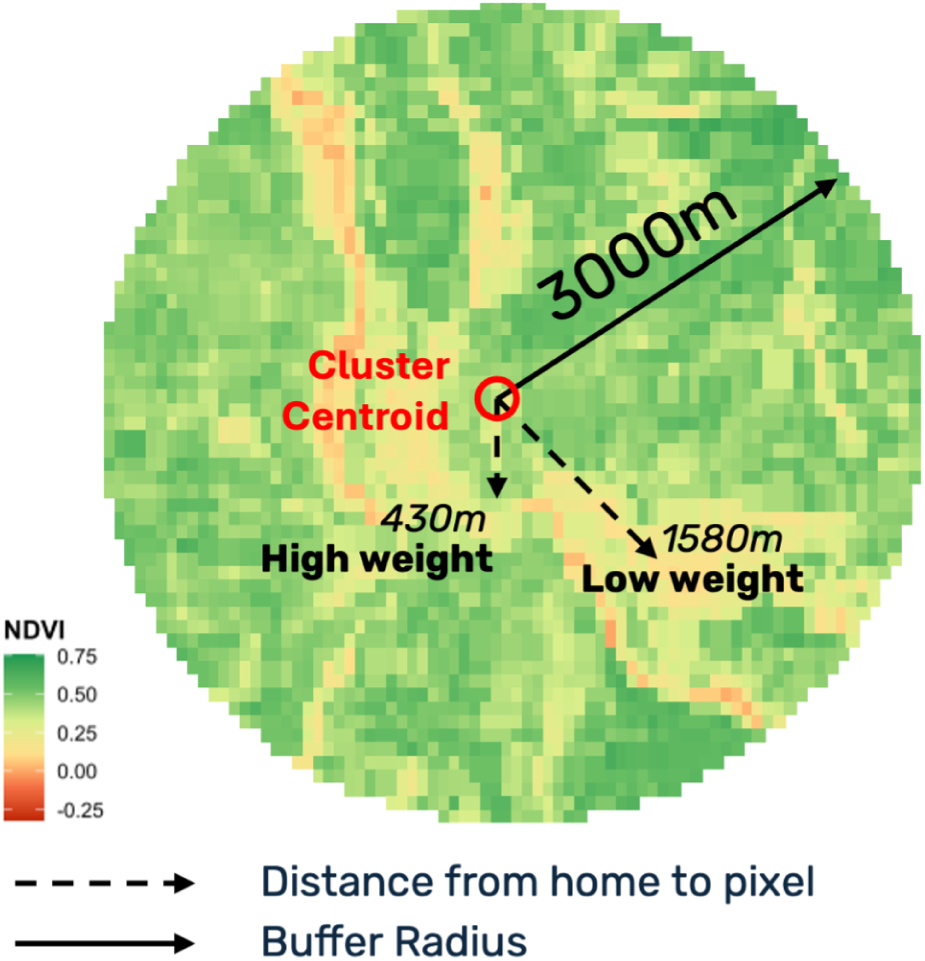
Illustration of the 3,000m NDVI buffer and distance-decay weighting scheme used in the human movement model. Higher weights are assigned to pixels near the DHS cluster centroid (“home”) and lower weights to pixels farther away.

Thus, the normalised weight for pixel *i* in the vicinity of child *j*’s home was defined as

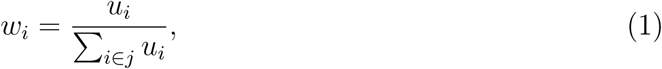

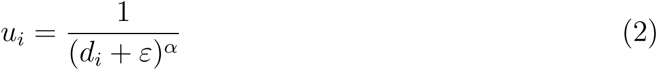

where *d_i_* is the distance from the home location to pixel *i* and *ε* is a small constant (10^−50^) included for numerical stability to prevent floating point errors. The decay parameter, *α*, controlled the rate at which weights declined with distance. Weights summed to 1 i.e. Σ*_i_*_∈_*_j_ w_i_* = 1 for each child.

Detailed mobility data (e.g., GPS trajectories or time–activity information) are often unavailable in population surveys such as the ADHS, although the methodology is intended to incorporate such data where available. The distance-decay function offers one tractable approach to representing movement in exposure-health studies without requiring detailed mobility data (Wei et al. 2023). More complex formulations, such as gravity models (Wheeler 2005), were not used as they rely on origin-destination data, which are difficult to define given that routine mobility may involve multiple destination types, such as homes, shops, and parks, each visited with different probabilities.

### 2.4 Statistical Methodology

We developed three structurally similar models that differ in their assumptions about the NDVI–ARI exposure–response relationship and movement-weighting structure (Table 1). The fixed-weight (FW) and learned-weight (LW) models assumed a linear NDVI-ARI association, whereas the restricted cubic spline (RCS) model allowed for nonlinear effects. All models used inverse-distance weighting to construct a movement-modelled exposure surface, with the decay parameter *α* either fixed (*α* = 1) or learned from the data. Estimating the decay parameter allowed the movement-weighting structure itself to be informed by the data, providing a data-driven representation of how nearby environments contribute to ARI risk.

**Table 1:** Summary of three model specifications.

| Model | NDVI-ARI Effect | Weights ($w_i$) | Decay Parameter ($\alpha$) |
| --- | --- | --- | --- |
| Fixed-weight (FW) | Linear | Fixed | $\alpha = 1$ |
| Learned-weight (LW) | Linear | Determined by $\alpha$ | $\alpha$ learned from data<br>Prior: $\log \alpha \sim \mathcal{N}(-1, 1)$ |
| RCS | Non-linear | Fixed | $\alpha = 1$ |

To estimate the association between pixel-level NDVI and ARI risk while incorporating an assumed movement pattern, the binary ARI outcome for child *j* was modelled as

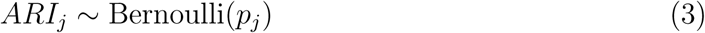

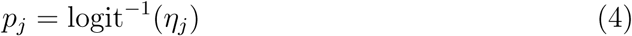

where *p_j_*denotes the probability of ARI for child *j*. The linear predictor for child *j*, *η_j_*, was defined separately for the three models. For the fixed-weight and learned-weight models,

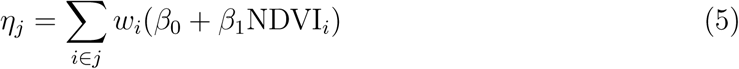

which represents a proximity-weighted sum of the NDVI values across all pixels within the buffer, with weights *w_i_* declining as distance from the home location increases.

For the RCS model,

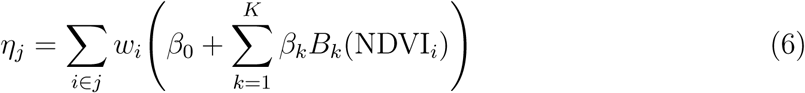

where *B_k_*(.) are the RCS basis functions and *K* is the number of spline terms. Restricted cubic spline basis functions were generated using the *splines* package in R. Knots were placed at the 10th, 50th, and 99th percentiles of the NDVI distribution, yielding two spline parameters *β*_1_ and *β*_2_.

Thus, each model estimated the probability of ARI as a function of the greenness surrounding the child’s home location, weighted according to how individuals are assumed to move within their local environment.

All models were fitted in a Bayesian framework. Uninformative priors were placed on the intercept and slope parameters (*β*_0_, *β*_1_ ~ *N* (0, 10^2^)). For the learned-weight model, a weakly informative prior was placed on the log-transformed decay parameter (log *α* ~ *N* (−1, 1^2^)), centring the prior near *α* = *e*^−1^ ≈ 0.37. All statistical models were implemented in C++ using the Template Model Builder (TMB) framework and interfaced through R. We elected to use TMB with Laplace approximation as it avoids the computational burden of full MCMC when modelling large numbers of individuals and pixels per individual. All computations were performed on the University of Leicester’s High-Performance Computing cluster (ALICE).

#### 2.4.1 Simulation Study

A simulation study was conducted to evaluate each model’s ability to recover parameter values under different sample sizes: the observed DHS dataset (N = 1,114) and two synthetic datasets created by sampling with replacement (N = 50,000 and N = 100,000). Figure 2 summarises the model development and testing workflow.

**Figure 2:**
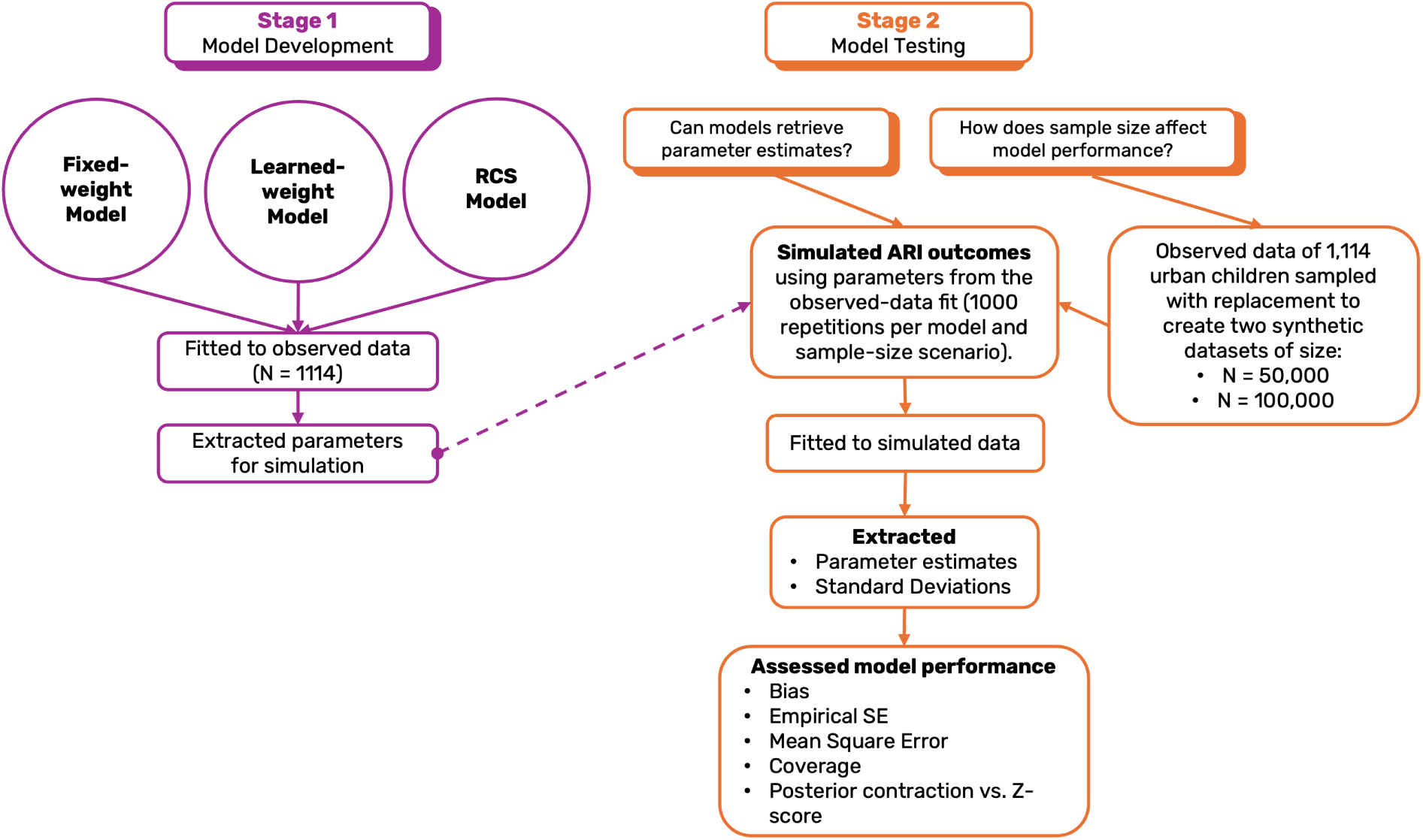
Overview of the model development (Stage 1) and model testing (Stage 2) workflow used to evaluate parameter recovery and sample-size effects.

To generate simulated outcomes, each model was first fitted to the observed DHS dataset to obtain baseline parameter estimates. The point values of these estimates were subsequently used as ground truth. Linear predictors and corresponding ARI probabilities were computed. Simulated ARI outcomes were drawn from Bernoulli distributions (*rbinom* in R) 1,000 times for every model and sample size scenario.

Model performance was assessed using bias, empirical standard error, mean squared error, and 95% credible interval coverage. Additionally, posterior contraction and posterior z-scores were visualised to understand the models’ ability to learn from the data (Betancourt 2020).

## 3 Results

### 3.1 Exploratory Analysis

Across the study population of 1,114 urban children in Albania, 23 (2.1%) were reported to have ARI symptoms. The spatial distribution of ARI cases, as shown in Figure 3, appear geographically dispersed, with no clear clustering of cases. The survey included 322 urban clusters with a median of 3 (IQR: 2-5) households per cluster and 3,740 (IQR: 3,737–3,743) NDVI pixels per household. Summary characteristics of the observed DHS data (N = 1,114) are provided in Table 2. No noticeable differences in NDVI exposure and the median distance from cluster centroid to surrounding pixels were found for children with and without ARI.

**Figure 3:**
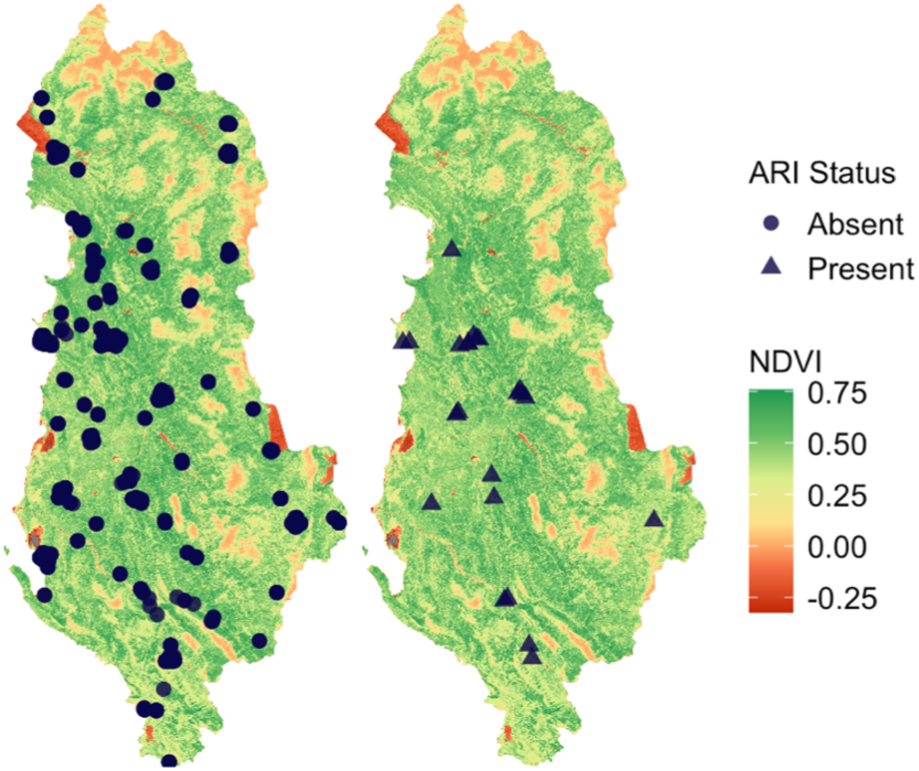
Spatial distribution of NDVI split by ARI cases in the 2017-2018 Albania DHS.

**Table 2:** Study Population (N = 1114). Summary statistics for individuals with ARI (N = 23) and without ARI (N = 1091). Total number of pixels: 4,166,554. Median and 25th to 75th percentile in parenthesis.

|  | ARI Present<br>(86,012 pixels) | ARI Absent<br>(4,080,542 pixels) |
| --- | --- | --- |
| NDVI | 0.41 (0.27–0.50) | 0.40 (0.25–0.49) |
| Distance from cluster centroid to pixel (m) | 2120 (1499–2597) | 2120 (1500–2597) |

To illustrate spatial heterogeneity in environmental exposure, Figure 4 presents three representative DHS cluster buffers. NDVI distributions varied notably both within and between buffers: some clusters were situated in predominantly green areas, whereas others encompassed low-NDVI coastal, agricultural, or built-up regions. These local spatial patterns also shaped the NDVI–distance relationship. In several clusters, NDVI increased slightly with distance from the centroid, highlighting that vegetation cover was neither uniform nor strictly monotonic with proximity to the home location.

**Figure 4:**
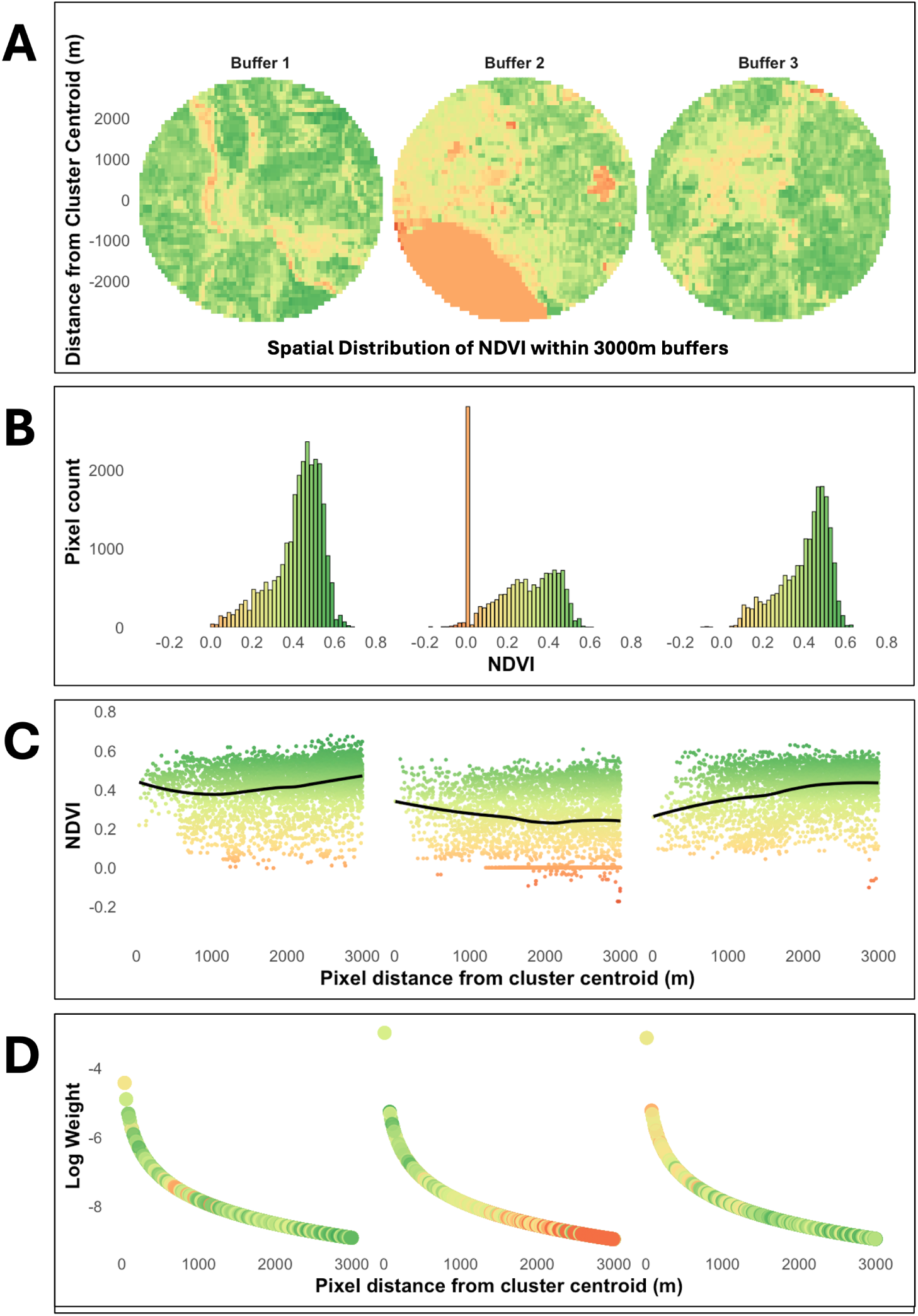
Spatial pattern of NDVI for three example buffers around DHS cluster centroids, selected to demonstrate the variability in environmental exposure within and between clusters. For each buffer, four complementary visualisations are presented: (A) a spatial map of pixel-level NDVI within the 3,000m buffer; (B) the distribution of NDVI values (including a spike at NDVI = 0 for Buffer 2. Because this cluster is located in a coastal area, a substantial portion of the buffer overlaps water or shoreline, where NDVI values are zero); (C) the relationship between NDVI and distance from cluster centroid; (D) the movement weights, based on inverse-distance weighting (α=1), fixed a priori as in the fixed-weight model. Weights were log-transformed to improve visualisation of the distance-decay pattern.

### 3.2 Fitting to Observed Data

We first fitted the fixed-, learned-, and RCS models to the observed DHS dataset (N = 1,114), retrieving the parameter values shown in Table 3. In both the fixed-weight and learned-weight models, the estimated linear effect of NDVI on ARI was positive but small, with credible intervals encompassing zero. These estimates were solely used as the data-generating parameters in the simulation study.

**Table 3:** Parameter estimates obtained from fitting each model to the observed data.

| Parameters | Fixed-weight | Learned-weight | RCS |
| --- | --- | --- | --- |
| $\log \alpha$ | | 0.09 | |
| $\beta_0$ | -4.30 | -5.27 | -5.52 |
| $\beta_1$ | 1.23 | 3.85 | 3.94 |
| $\beta_2$ | | | -0.19 |

In the learned-weight model, the estimated decay parameter, *α*, was *>* 1, which corresponds mathematically to a sharper down-weighting of pixels farther from the home location. If *α* were *<* 1, the weighting structure would approach a more uniform surface (similar to taking a simple mean across the buffer), but this did not occur in the observed-data fit.

### 3.3 Fitting to Simulated Data

We summarise parameter recovery and sample-size performance across the three model specifications in Supplementary Table **??**, reporting bias, empirical standard error, mean squared error, 95% credible interval coverage and the Monte Carlo Error for each measure and modelling scenario.

#### Fixed-weight Model

The fixed-weight model improved in parameter recovery as sample size increased (Figure 5). For both parameters *β*_0_ and *β*_1_, bias decreased from 0.231 and −0.787 at N = 1,114 to 0.006 and −0.017 at N = 100,000, respectively. We saw corresponding reductions in EmpSE and MSE. For example, the EmpSE for *β*_1_ declined from 2.055 at N = 1,114 to 0.225 at N = 100,000, while MSE decreased from 4.840 to 0.051. Coverage remained stable near the nominal 95% level across larger sample sizes (Figure 6).

**Figure 5:**
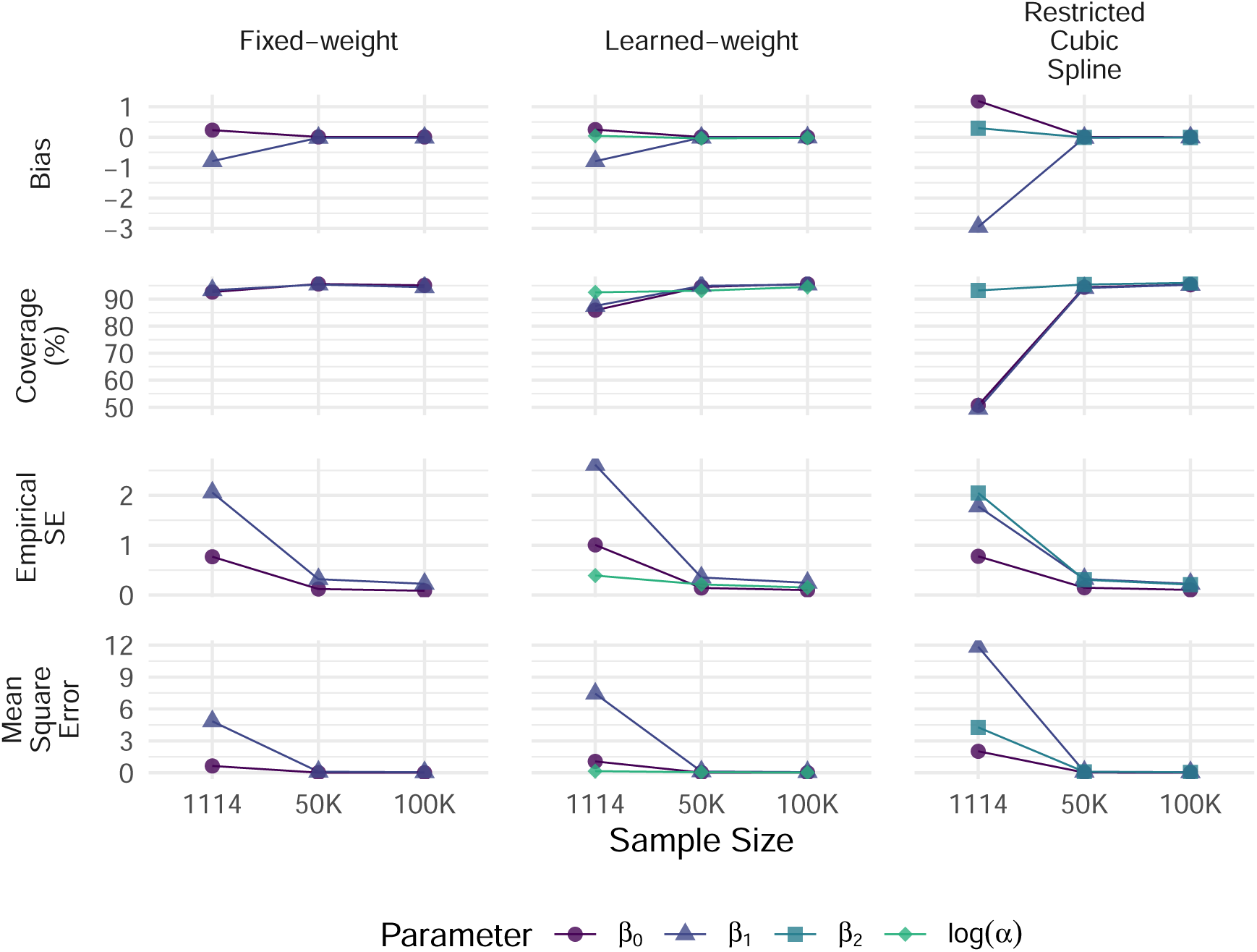
Performance measures across different sample size and modelling scenarios. As sample size increases, model performance improves across all three models: fixed-weight, learned-weight and restricted cubic spline.

**Figure 6:**
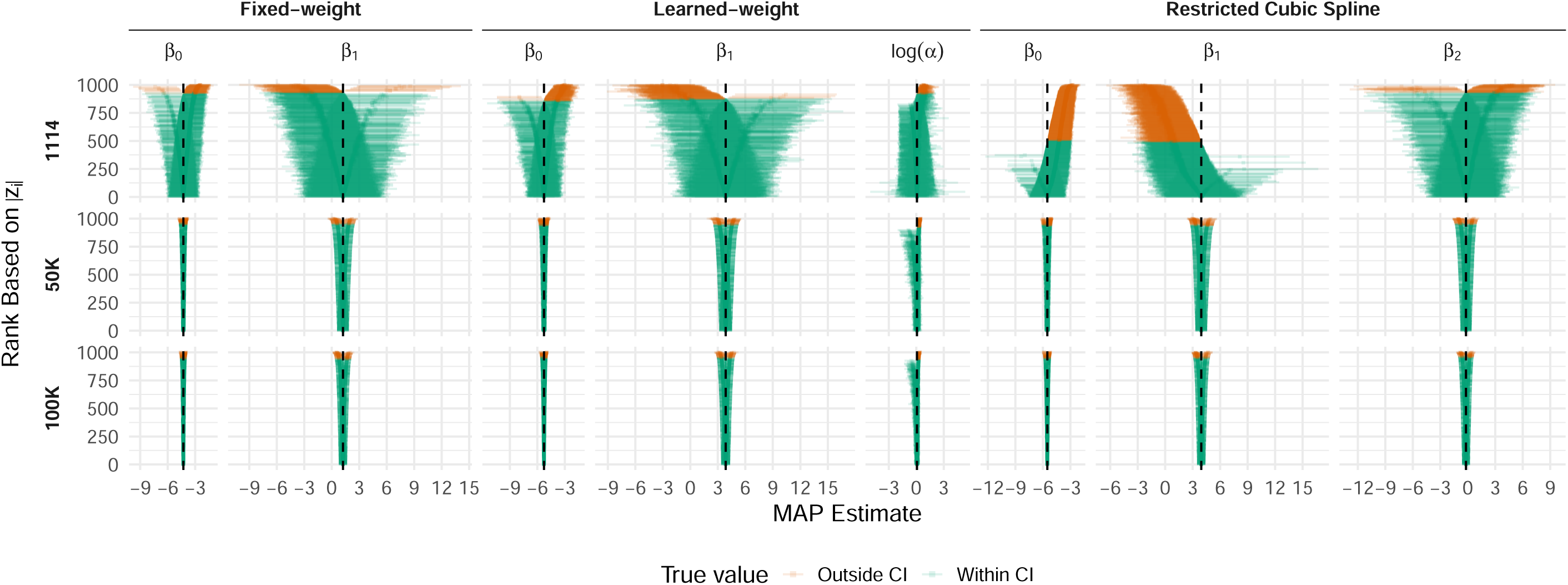
Ranked 95% credible intervals (zipper plots) for each parameter across three models and sample sizes. Each horizontal line represents one simulation, ranked by absolute posterior z-score (|z_i_|); the dashed vertical line marks the true parameter value. Green intervals contain the true value; orange intervals do not. At N = 1,114, intervals are wide and poorly centred, with substantial miscoverage—most notably for β_0_ and β_1_ in the RCS model. At N = 50,000 and N = 100,000, intervals narrow and converge tightly around the true value, reflecting the improvement in coverage and precision with increasing sample size.

At N = 1,114, simulations with higher posterior contraction for *β*_0_ also tended to exhibit negative posterior z-scores, indicating that the model continued to underestimate the NDVI effect even when the parameter was strongly informed by the data (Figure 7).

**Figure 7:**
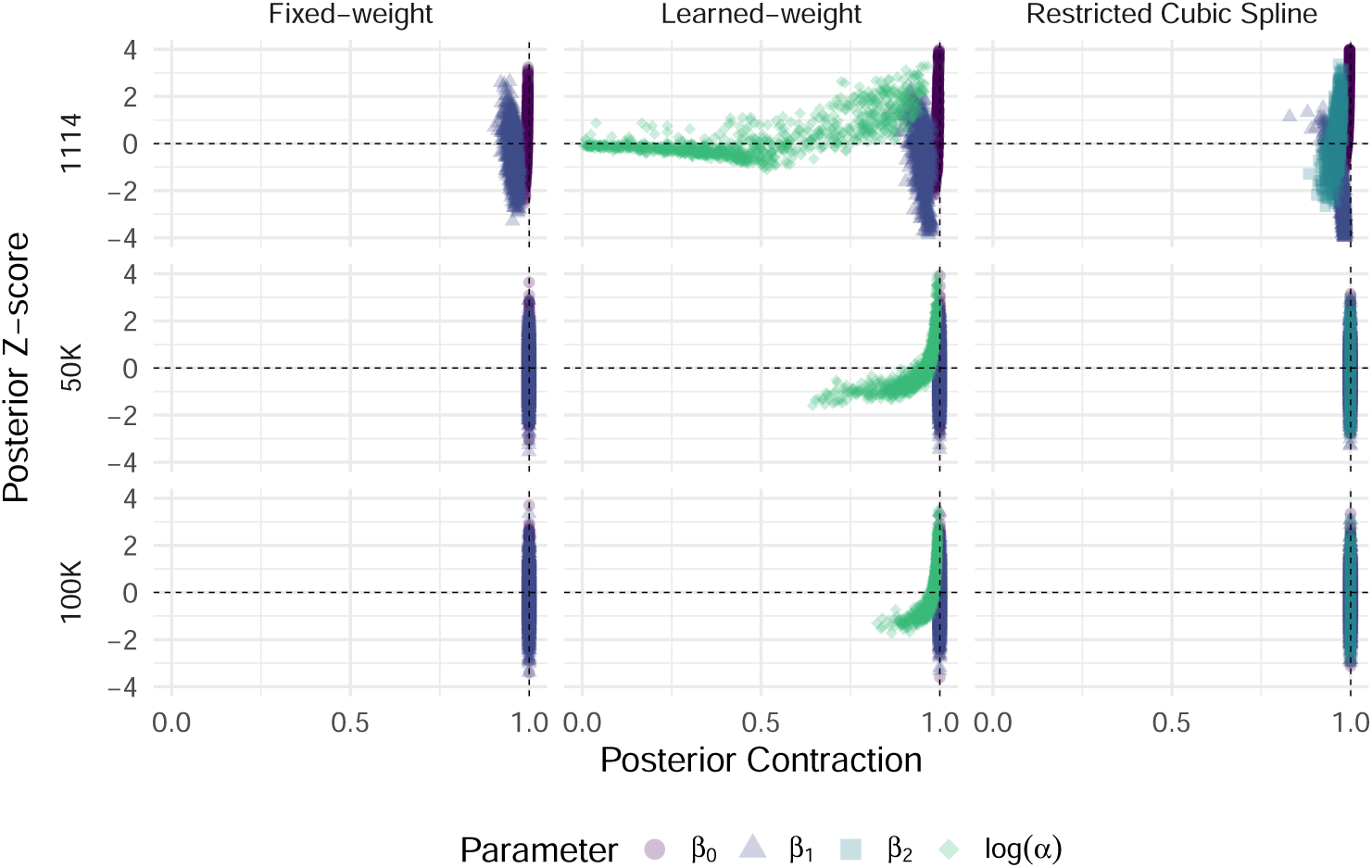
Posterior Contraction vs Z-score by model and sample size. Although log α improved as sample size increased, at N = 1114, the ‘j’-shaped pattern observed for log α, showed posterior z-scores close to zero when posterior contraction was low. This suggests that in some simulations the posterior differed little from the prior i.e. the model learned little additional information about α, resulting in MAP estimates that stayed close to the prior distribution, which was centred near the observed parameter value. Thus, the weights are largely determined by the prior, not the data at this sample size.

#### Learned-weight Model

The learned-weight model jointly estimated the NDVI–ARI association and the decay parameter *α*, which derives the pixel-level weights as demonstrated in Figure 8. By estimating *α* from the health outcome data, the model learns the spatial importance of NDVI pixels — that is, how rapidly the influence of NDVI pixels on ARI risk declines with distance from the proxy home location. For all parameters *β*_0_, *β*_1_, and log *α*, performance improved with larger sample sizes (Figure 5). For example, bias and EmpSE in the NDVI effect parameter *β*_1_ reduced from −0.790 and 2.612 at N = 1,114 to −0.011 and 0.245 at N = 100,000, respectively. Coverage for *β*_0_ and *β*_1_ fell below nominal levels (85.9% and 87.5%, respectively) at N = 1,114, but recovered to approximately 95% by N = 50,000. Coverage for log *α* remained close to nominal across all sample sizes (92.5%, 93.1%, and 94.5% at N = 1,114, 50,000, and 100,000).

**Figure 8:**
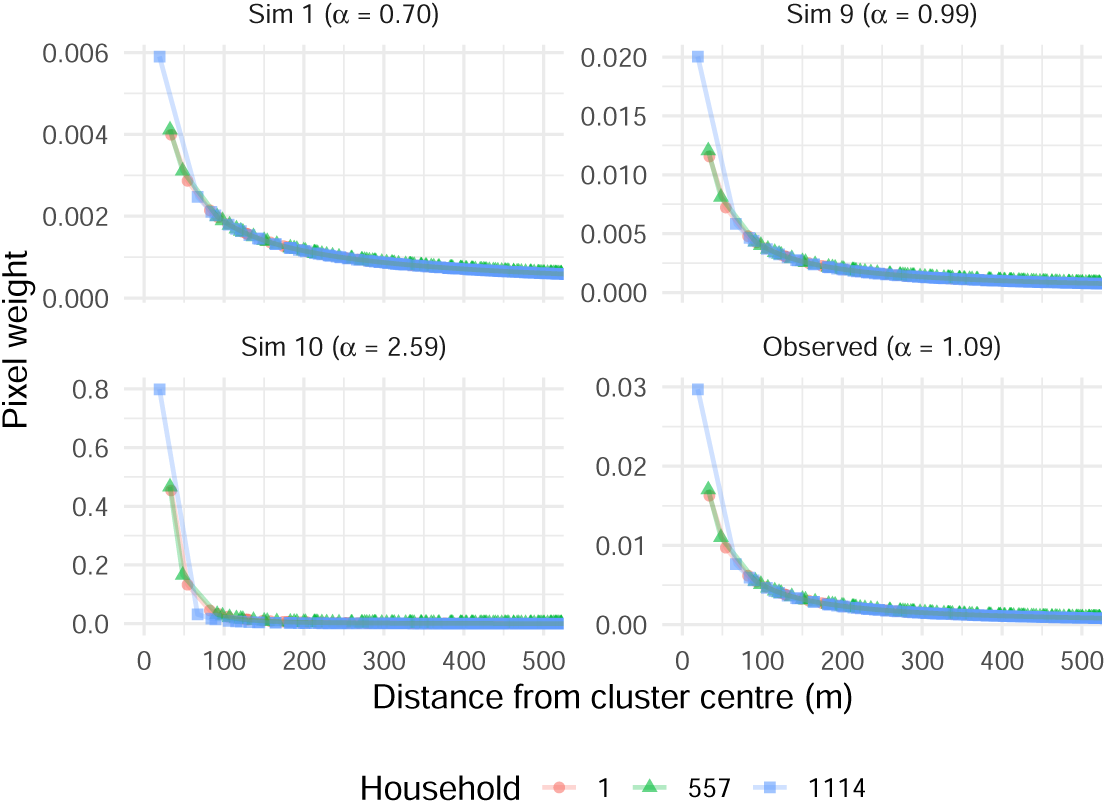
Pixel-level weights derived from learned α, zoomed to 0−500m from DHS cluster centroid for clarity. Random three simulations 1, 9 and 10 demonstrate how quickly the pixel-level weights decay with distance for three households (1, 557 and 1114) in the DHS dataset. Small values of α (0.70), show a steadier decline so that even pixels at 500m still contribute to health outcome, whilst large values of α (2.59), show that only pixels within a 100m locale matter most.

The learned decay parameter log *α* showed poor identifiability at N = 1,114, with weak posterior contraction (Figure 7). Although contraction improved at N = 100,000, a distinctive contraction–z-score pattern persisted. This reflects that identifiability depends not only on sample size but also on the spatial information of the NDVI data: when NDVI varies little with distance from the home location, different *α* values generate nearly identical weighted exposures, leaving the data unable to distinguish them.

#### RCS Model

The RCS model tested a non-linear relationship between the exposure and outcome. Like the fixed-weight and learned-weight models, parameter recovery was poor at N = 1,114. Highly biased and uncertain estimates were observed in *β*_0_ (bias = 1.190; EmpSE =0.777), *β*_1_ (bias = −2.948; EmpSE =1.777) and *β*_2_ (bias = 0.297; EmpSE =2.046). 95% credible interval coverage was only 50.7% and 49.4% for *β*_0_ and *β*_1_, respectively—far below the nominal 95% level (Figure 6). The nonlinear spline coefficient *β*_2_ showed different behaviour at N = 1,114 with coverage at 93.2%—near nominal, capturing the non-linear relationship in the interior of the NDVI distribution, where there are more data, and therefore was consequently better identified even at N = 1,114 (Figure 7).

Parameter recovery improved substantially at larger sample sizes. At N = 50,000, bias was negligible across all three parameters (*β*_0_ = 0.006, *β*_1_ = −0.019, *β*_2_ = −0.010), EmpSE values fell, and coverage was near nominal (94.4%, 94.1%, and 95.4%, respectively). This pattern was maintained at N = 100,000, where all parameters showed near-zero bias, further reduced EmpSE values (0.103, 0.225, and 0.209 for *β*_0_, *β*_1_, and *β*_2_), and coverage of 95.3%, 95.3%, and 96.0%.

## 4 Discussion

This study developed and evaluated three structurally similar statistical models in which pixel-level environmental data were weighted using a simple movement model to better represent environmental influences on health risk. Using ARI and greenness in Albania as an exemplar, we assessed each model’s ability to recover parameters under varying sample sizes and model complexities, including linear versus nonlinear exposure–response structures and fixed versus learned movement weights.

### 4.1 Disseminating Models

Given the many-to-one mapping of pixels to individuals, a potential source of spatial noise, and the variation in pixel counts across children, it was unsurprising that larger sample sizes (50,000 and 100,000 vs. 1,114) yielded substantially better parameter recovery and stability across all models. In practice, large-scale epidemiological cohorts, such as UK Biobank (Sudlow et al. 2015), CPRD (Herrett et al. 2015), and other national health surveys are increasingly available and offer sample sizes capable of supporting these exposure–health models. However, integrating these datasets with individual-level human movement data and fine-scale environmental exposures remains challenging due to the separate administrative systems in which these data are kept, privacy constraints, anonymisation requirements, and ethical considerations around linking geospatial information. Until more robust frameworks for linking health, environmental, and human mobility data become available, these statistical models may be applied at coarser spatial scales (e.g. census tracts or area-level analyses), where privacy protections are easier to maintain, or with spatial distribution-driven assumptions of where people are likely to spend their time as a proxy for mobility.

In keeping with the methodological focus of this work, we sought to keep our initial tests intentionally simple so as to isolate the behaviour of the statistical method itself in a controlled setting. For this reason, we employed a simple distance-decay model as a representation of local mobility. In practice, this method is not limited to this simple representation of human movement and does not require applying residential-based buffers.

The fixed-weight model, structurally the simplest, was both computationally efficient and robust at larger sample sizes. Its reliance on externally defined movement weights also makes it useful for applied settings where weights can be specified from prior studies, behavioural assumptions, or mobility data.

The learned-weight model simultaneously estimates the exposure–response relationship and the spatial influence of surrounding environments on individual risk. It performed poorly at smaller sample sizes, reflecting the inherent difficulty of learning latent spatial parameters from sparse outcome data. However, as sample size increased, the model recovered the decay parameter and exposure effects more reliably, demonstrating that with adequate data, it is possible to infer complex exposure patterns directly from the health data. By learning environmental influence from the data, the learned-weight model offers an alternative to the flat, fixed-radius buffers that dominate the exposure–health literature (Feleszko et al. 2024; Fuertes et al. 2020; Squillacioti et al. 2024), because it does not require strong, arbitrary assumptions about environmental influence. In contrast, conventional buffer approaches (whether 300 m, 1 km, or 3 km) implicitly assume uniform exposure within the buffer and zero exposure beyond it, despite well-documented evidence that estimated associations are sensitive to the choice of buffer size and shape (Browning and Lee 2017; Hartley et al. 2020; Tang et al. 2023). These arbitrary design decisions can alter effect estimates, contribute to exposure misclassification and, due to the heterogeneous buffer choices, hinder causal inference and evidence synthesis—a problem repeatedly highlighted in studies of buffer size, buffer type, and spatial resolution of exposure surfaces (Zare Sakhvidi et al. 2025; Jimenez et al. 2022; Kim and Kwan 2019). Our method addresses this buffer selection problem by allowing the data to inform how exposure influence accumulates across space. This framework can be applied in two distinct ways: to understand how the spatial properties of an environmental surface influence health at varying distances from a reference location, without requiring individual movement data; or, where movement data are available, to incorporate individuals’ actual locations and time spent across space, enabling direct assessment of personal environmental interaction. In either application, this is particularly valuable for identifying which disaggregated locations contribute most strongly to elevated or reduced risk and therefore has clear implications for targeted environmental interventions or policy.

The RCS model captured a nonlinear relationship between greenness and ARI. While the model recovered key parameters and—like the others—performed best with approximately 100,000 individuals, the boundary spline term proved difficult to estimate. This is likely due to the combination of sparse NDVI data at the extremes of the distribution and the rarity of ARI in the observed dataset, leading to weak identification of spline behaviour at the tails. These challenges are consistent with well-documented issues in spline estimation under limited information (Gauthier et al. 2020).

### 4.2 Strengths

These models address the difficulty of linking fine-resolution environmental data to individual health outcomes. Our results show that, despite limitations in mobility data, spatial misalignment, and the computational demands of high-resolution exposure modelling, a simple movement representation can be integrated into epidemiological analyses without resorting to averaged exposure metrics. By allowing environmental influence to vary with how individuals interact with their surroundings (e.g., spending more time near home), the framework moves toward reducing the exposure misclassification inherent in residential-only approaches.

A further key strength of the modelling framework is its flexibility. The structure is not specific to NDVI or ARI: NDVI could be replaced with PM_2.5_, temperature, noise, ultraviolet radiation, or other environmental surfaces, and ARI could be replaced with a wide range of health outcomes, including asthma, COPD, mental health indicators, or cardiovascular events. The models accommodate linear or nonlinear exposure–response relationships and can use either externally specified movement weights or weights learned directly from the data.

Finally, the models remain interpretable: the exposure effect is still expressed as a change in the log-odds of disease, but its influence varies spatially through the weights.

### 4.3 Limitations

Several modelling simplifications were made for parsimony in this work, particularly to reduce the computational burden of developing and testing these models. These include the use of a simplified movement model that does not fully capture real-world mobility patterns; the reliance on DHS cluster centroids as proxies for household locations, which introduces potential exposure misclassification; and the focus on a single exposure (NDVI) despite the known multifactorial determinants of ARI risk. Additionally, choices regarding priors, buffer size, and spatial or temporal NDVI resolution are all factors known to influence results, though exploring these sensitivities was beyond the scope of this methodological work as this was an example case study. Future work will apply this framework to an applied study, addressing epidemiological questions on the influence of environmental exposures on health whilst accounting for human movement.

These models were also computationally intensive. Unsurprisingly, both the learned-weight and RCS models were more computationally demanding than the fixed-weight model, reflecting the added complexity of estimating latent decay parameters (e.g., log *α*) and nonlinear exposure–response functions. Across the 100,000-individual simulation, approximately 340 million NDVI pixels were processed, and a single learned-weight model required around 40 minutes to run. To keep the models feasible to run, we used the Laplace approximation implemented in TMB, which provides an efficient way to fit complex models without the full computational cost of Bayesian sampling methods. Although this approach allowed us to manage the demands of the current framework, incorporating more detailed movement models or time-varying exposure surfaces would still introduce substantial additional computational challenges. This underscores the need to balance model complexity with data resolution to ensure computational feasibility and efficiency.

### 4.4 Comparison with Literature

Recent work in disease modelling has also used human movement to refine exposure estimates. Knoblauch et al. (2025) developed an intraday *Aedes*–human exposure model that combines hourly mobility patterns with mosquito biting activity to generate time-specific estimates of bite risk based on where people move over the course of a day. Mills et al. (2025) similarly incorporated mobility into a Bayesian spatiotemporal model of dengue transmission, using movement flows between neighbourhoods to shape both local force of infection and the spread of cases. Sugg et al. (2019) measured personal ambient temperature using wearable sensors, then compared it with fixed-site weather station data, showing only a weak relationship and warning against relying on ambient stations alone for health-effect studies. These studies are examples of replacing buffer-based risk with movement-informed exposure (where people actually are, when it matters). The studies share conceptual similarities with our framework, particularly the idea of weighting risk according to where individuals are located in space or time. Knoblauch et al. (2025) and Mills et al. (2025) remain focused on mosquito ecology and infectious disease dynamics, while Sugg et al. (2019) weighted thermal exposure by each worker’s own spatiotemporal exposure captured via sensors. In contrast, our model applies spatial weighting to satellite-derived greenness and links these weighted surfaces directly to individual-level respiratory health outcomes, offering a different public health context.

Our methodological approach extends the commonly used buffer-based methods in environmental epidemiology (Browning and Lee 2017; Zare Sakhvidi et al. 2025) because it does not treat all locations inside the buffer as equally important. Instead, it allows the influence of environmental exposures to vary depending on how individuals interact with space. This work contributes to the environmental exposure–health literature by providing a methodological alternative to fixed-buffer exposure assignment and by addressing long-standing concerns about buffer sensitivity (Zare Sakhvidi et al. 2025) and exposure misclassification.

## 5 Conclusion

This study introduced a flexible approach for incorporating simple movement patterns into models linking fine-scale environmental data with individual health outcomes. Using pixel-level NDVI and proximity-based weights, we demonstrated through a simulation study that these models can recover underlying parameters at large sample sizes and help address exposure misclassification arising from residential-only metrics. This framework simultaneously estimates the exposure-response relationship and learns the spatial influence of disaggregated, pixel-level environmental data directly from health outcome data.

Although intentionally simplified in this study, this methodological framework re-mains scalable, interpretable, and adaptable to other exposures and outcomes, offering a foundation for future work that integrates richer mobility-informed exposure-health research.

## Code and Data Availability

The statistical models, simulation code, and analysis scripts used in this study are publicly available on GitHub at https://github.com/hadiqa-tahir/exposure-disaggregation-regression. The DHS data used in the case study are available upon application from the DHS Programme (https://dhsprogram.com).

## CRediT Authorship Contribution Statement

**Hadiqa Tahir:** Conceptualization, Methodology, Software, Formal Analysis, Investigation, Data Curation, Writing – Original Draft, Visualization, Project Administration; **Simon Smart:** Resources, Writing – Review & Editing; **Samuel Cai:** Conceptualization, Resources; **André Ng:** Supervision; **Joshua Vande Hey:** Conceptualization, Writing – Review & Editing, Supervision; **Tim CD Lucas:** Conceptualization, Methodology, Investigation, Data Curation, Validation, Resources, Funding Acquisition, Project Administration, Supervision, Writing – Review & Editing

## References

Banerjee, Sudipto, Bradley P. Carlin, Alan E. Gelfand, and Sudipto Banerjee (Dec. 17, 2003). Hierarchical Modeling and Analysis for Spatial Data. New York: Chapman and Hall/CRC. 472 pp. isbn: 978-0-429-20523-1. doi: 10.1201/9780203487808.

Betancourt, Michael (Apr. 1, 2020). *Towards A Principled Bayesian Workflow*. url: https://betanalpha.github.io/assets/case_studies/principled_bayesian_workflow.html (visited on 02/20/2026).

Browning, Matthew and Kangjae Lee (June 23, 2017). “Within What Distance Does “Greenness” Best Predict Physical Health? A Systematic Review of Articles with GIS Buffer Analyses across the Lifespan”. In: International Journal of Environmental Research and Public Health 14.7, p. 675. issn: 1660-4601. doi: 10.3390/ijerph14070675. PMID: 28644420.

Chandia-Poblete, Damian, Thomas Cole-Hunter, Melissa Haswell, and Kristiann C. Heesch (Dec. 1, 2022). “The Influence of Air Pollution Exposure on the Short- and Long-Term Health Benefits Associated with Active Mobility: A Systematic Review”. In: Science of The Total Environment 850, p. 157978. issn: 0048-9697. doi: 10.1016/j.scitotenv.2022.157978. url: https://www.sciencedirect.com/science/article/pii/S004896972205077X (visited on 12/04/2025).

Chen, Edith et al. (Apr. 1, 2017). “Difficult Family Relationships, Residential Greenspace, and Childhood Asthma”. In: Pediatrics 139.4, e20163056. issn: 0031-4005. doi: 10.1542/peds.2016-3056. url: https://doi.org/10.1542/peds.2016-3056 (visited on 06/26/2025).

Citron, Daniel T, Shankar Iyer, Robert C Reiner, and David L Smith (2021). “Activity Space Maps: A Novel Human Mobility Data Set for Quantifying Time Spent at Risk”. In: medRxiv, pp. 2021–08.

Feleszko, Wojciech et al. (2024). “Early-Life Exposure to Residential Greenness and Risk of Asthma in a U.S. Bronchiolitis Cohort”. In: Allergy 79.11, pp. 3036–3046. issn: 1398-9995. doi: 10.1111/all.16359. url: https://onlinelibrary.wiley.com/doi/abs/10.1111/all.16359 (visited on 07/11/2025).

Fernandes, Amanda et al. (Oct. 1, 2023). “Availability, Accessibility, and Use of Green Spaces and Cognitive Development in Primary School Children”. In: Environmental Pollution 334, p. 122143. issn: 0269-7491. doi: 10.1016/j.envpol.2023.122143. url: https://www.sciencedirect.com/science/article/pii/S0269749123011454 (visited on 11/28/2025).

Fuertes, Elaine et al. (2020). “Residential Greenspace and Lung Function up to 24 Years of Age: The ALSPAC Birth Cohort”. In: Environment international 140, p. 105749. issn: 0160-4120.

Gauthier, J., Q. V. Wu, and T. A. Gooley (Apr. 1, 2020). “Cubic Splines to Model Relationships between Continuous Variables and Outcomes: A Guide for Clinicians”. In: Bone Marrow Transplantation 55.4, pp. 675–680. issn: 0268-3369, 1476-5365. doi: 10.1038/s41409-019-0679-x. url: https://www.nature.com/articles/s41409-019-0679-x (visited on 06/08/2026).

Hartley, Kim, Patrick Ryan, Cole Brokamp, and Gordon L. Gillespie (May 1, 2020). “Effect of Greenness on Asthma in Children: A Systematic Review”. In: Public Health Nursing 37.3, pp. 453–460. issn: 1525-1446. doi: 10.1111/phn.12701. url: https://onlinelibrary.wiley.com/doi/10.1111/phn.12701 (visited on 06/26/2025).

Herrett, Emily et al. (June 2015). “Data Resource Profile: Clinical Practice Research Datalink (CPRD)”. In: International Journal of Epidemiology 44.3, pp. 827–836. issn: 0300-5771. doi: 10.1093/ije/dyv098. PMID: 26050254. url: https://pmc.ncbi.nlm.nih.gov/articles/PMC4521131/ (visited on 06/08/2026).

Idrose, Nur Sabrina, Caroline J. Lodge, Bircan Erbas, Jo A. Douglass, Dinh S. Bui, and Shyamali C. Dharmage (Jan. 2022). “A Review of the Respiratory Health Burden Attributable to Short-Term Exposure to Pollen”. In: International Journal of Environmental Research and Public Health 19.12, p. 7541. issn: 1660-4601. doi: 10.3390/ijerph19127541. url: https://www.mdpi.com/1660-4601/19/12/7541 (visited on 12/05/2025).

Institute of Statistics, Institute of Public Health, and ICF (Oct. 1, 2018). *Albania Demographic and Health Survey 2017-18*. url: https://dhsprogram.com/publications/publication-FR348-DHS-Final-Reports.cfm (visited on 06/10/2026).

Jimenez, Raquel B., Kevin J. Lane, Lucy R. Hutyra, and M. Patricia Fabian (Mar. 2022). “Spatial Resolution of Normalized Difference Vegetation Index and Greenness Exposure Misclassification in an Urban Cohort”. In: Journal of Exposure Science & Environmental Epidemiology 32.2, pp. 213–222. issn: 1559-064X. doi: 10.1038/s41370-022-00409-w. PMID: 35094014.

Johannessen, Ane, Shanshan Xu, Achenyo Peace Abbah, and Christer Janson (June 2023). “Greenness Exposure: Beneficial but Multidimensional”. In: Breathe (Sheffield, England) 19.2, p. 220221. issn: 1810-6838. doi: 10.1183/20734735.0221-2022. PMID: 37645023.

Kim, Junghwan and Mei-Po Kwan (Jan. 2019). “Beyond Commuting: Ignoring Individuals’ Activity-Travel Patterns May Lead to Inaccurate Assessments of Their Exposure to Traffic Congestion”. In: International Journal of Environmental Research and Public Health 16.1, p. 89. issn: 1660-4601. doi: 10.3390/ijerph16010089. url: https://www.mdpi.com/1660-4601/16/1/89 (visited on 03/12/2026).

Kim, Sunghyub, Athanasios Damialis, Athanasios Charalampopoulos, Dayne H. Voelker, and Andrew C. Rorie (Feb. 1, 2025). “The Effect of Climate Change on Allergen and Irritant Exposure”. In: The Journal of Allergy and Clinical Immunology: In Practice 13.2, pp. 266–273. issn: 2213-2198, 2213-2201. doi: 10.1016/j.jaip.2024.12.019. PMID: 39710224. url: https://www.jaci-inpractice.org/article/S2213-2198(24)01262-5/fulltext (visited on 06/26/2025).

Knoblauch, Steffen et al. (Mar. 7, 2025). “Modeling Intraday Aedes-human Exposure Dynamics Enhances Dengue Risk Prediction”. In: Scientific Reports 15.1, p. 7994. issn: 2045-2322. doi: 10.1038/s41598-025-91950-9. url: https://www.nature.com/articles/s41598-025-91950-9 (visited on 06/29/2025).

Kwan, Mei-Po (Sept. 1, 2012). “The Uncertain Geographic Context Problem”. In: Annals of the Association of American Geographers 102.5, pp. 958–968. issn: 0004-5608. doi: 10.1080/00045608.2012.687349. url: https://doi.org/10.1080/00045608.2012.687349 (visited on 06/24/2025).

Kwan, Mei-Po (2018). “The Neighborhood Effect Averaging Problem (NEAP): An Elusive Confounder of the Neighborhood Effect”. In: International journal of environmental research and public health 15.9, p. 1841. issn: 1660-4601.

Letellier, Noémie et al. (June 2022). “Air Pollution and Metabolic Disorders: Dynamic versus Static Measures of Exposure among Hispanics/Latinos and Non-Hispanics”. In: Environmental Research 209, p. 112846. issn: 1096-0953. doi: 10.1016/j.envres.2022.112846. PMID: 35120894.

Li, Zhenlong, Huan Ning, Fengrui Jing, and M. Naser Lessani (Jan. 19, 2024). “Understanding the Bias of Mobile Location Data across Spatial Scales and over Time: A Comprehensive Analysis of SafeGraph Data in the United States”. In: PLOS ONE 19.1, e0294430. issn: 1932-6203. doi: 10.1371/journal.pone.0294430. url: https://journals.plos.org/plosone/article?id=10.1371/journal.pone.0294430 (visited on 03/13/2026).

Lu, Yougeng (Oct. 1, 2021). “Beyond Air Pollution at Home: Assessment of Personal Exposure to PM2.5 Using Activity-Based Travel Demand Model and Low-Cost Air Sensor Network Data”. In: Environmental Research 201, p. 111549. issn: 0013-9351. doi: 10.1016/j.envres.2021.111549. url: https://www.sciencedirect.com/science/article/pii/S0013935121008434 (visited on 06/24/2025).

Ma, Jing, Yinhua Tao, Mei-Po Kwan, and Yanwei and Chai (Mar. 3, 2020). “Assessing Mobility-Based Real-Time Air Pollution Exposure in Space and Time Using Smart Sensors and GPS Trajectories in Beijing”. In: Annals of the American Association of Geographers 110.2, pp. 434–448. issn: 2469-4452. doi: 10.1080/24694452.2019.1653752. url: https://doi.org/10.1080/24694452.2019.1653752 (visited on 06/29/2025).

Mills, Cathal et al. (June 4, 2025). *The Time- and Space-Varying Roles of Human Mobility in Shaping Urban Dengue Epidemics*. doi: 10.1101/2025.06.03.25328731. url: https://www.medrxiv.org/content/10.1101/2025.06.03.25328731v1 (visited on 06/29/2025). Pre-published.

Nieuwenhuijsen, Mark J. et al. (Apr. 1, 2014). “Positive Health Effects of the Natural Outdoor Environment in Typical Populations in Different Regions in Europe (PHENOTYPE): A Study Programme Protocol”. In: BMJ Open 4.4, e004951. issn: 2044-6055, 2044-6055. doi: 10.1136/bmjopen-2014-004951. PMID: 24740979. url: https://bmjopen.bmj.com/content/4/4/e004951 (visited on 06/26/2025).

Oliver, Lisa N, Nadine Schuurman, and Alexander W Hall (2007). “Comparing Circular and Network Buffers to Examine the Influence of Land Use on Walking for Leisure and Errands”. In: International journal of health geographics 6, pp. 1–11.

Poorthuis, Ate, Qingqing Chen, and Matthew Zook (Nov. 1, 2024). “A Nationwide Dataset of De-Identified Activity Spaces Derived from Geotagged Social Media Data”. In: Environment and Planning B: Urban Analytics and City Science 51.9, pp. 2264–2275. issn: 2399-8083. doi: 10.1177/23998083241264051. url: https://doi.org/10.1177/23998083241264051 (visited on 03/12/2026).

Smith, James David et al. (Nov. 1, 2016). “London Hybrid Exposure Model: Improving Human Exposure Estimates to NO2 and PM2.5 in an Urban Setting”. In: Environmental Science & Technology 50.21, pp. 11760–11768. issn: 0013-936X. doi: 10.1021/acs.est.6b01817. url: https://doi.org/10.1021/acs.est.6b01817 (visited on 03/13/2026).

Squillacioti, Giulia et al. (Apr. 15, 2024). “Different Greenness Exposure in Europe and Respiratory Outcomes in Youths. A Systematic Review and Meta-Analysis”. In: Environmental Research 247, p. 118166. issn: 0013-9351. doi: 10.1016/j.envres.2024.118166. url: https://www.sciencedirect.com/science/article/pii/S0013935124000707 (visited on 06/26/2025).

Su, Jason G. et al. (July 2024). “Health Effects of Air Pollution on Respiratory Symptoms: A Longitudinal Study Using Digital Health Sensors”. In: Environment International 189, p. 108810. issn: 01604120. doi: 10.1016/j.envint.2024.108810. url: https://linkinghub.elsevier.com/retrieve/pii/S0160412024003969 (visited on 12/05/2025).

Sudlow, Cathie et al. (Mar. 31, 2015). “UK Biobank: An Open Access Resource for Identifying the Causes of a Wide Range of Complex Diseases of Middle and Old Age”. In: PLOS Medicine 12.3, e1001779. issn: 1549-1676. doi: 10.1371/journal.pmed.1001779. url: https://journals.plos.org/plosmedicine/article?id=10.1371/journal.pmed.1001779 (visited on 06/08/2026).

Sugg, Margaret M., Scott Stevens, and Jennifer D. Runkle (June 2019). “Estimating Personal Ambient Temperature in Moderately Cold Environments for Occupationally Exposed Populations”. In: Environmental Research 173, pp. 497–507. issn: 00139351. doi: 10.1016/j.envres.2019.03.066. url: https://linkinghub.elsevier.com/retrieve/pii/S0013935119302002 (visited on 12/05/2025).

Tang, Mingcheng, Wei Liu, Haifang Li, and Fengyi Li (Dec. 6, 2023). “Greenness and Chronic Respiratory Health Issues: A Systematic Review and Meta-Analysis”. In: Frontiers in Public Health 11. issn: 2296-2565. doi: 10.3389/fpubh.2023.1279322. url: https://www.frontiersin.org/journals/public-health/articles/10.3389/fpubh.2023.1279322/full (visited on 06/26/2025).

Tayarani, Mohammad and Gregory Rowangould (Mar. 1, 2020). “Estimating Exposure to Fine Particulate Matter Emissions from Vehicle Traffic: Exposure Misclassification and Daily Activity Patterns in a Large, Sprawling Region”. In: Environmental Research 182, p. 108999. issn: 0013-9351. doi: 10.1016/j.envres.2019.108999. url: https://www.sciencedirect.com/science/article/pii/S0013935119307960 (visited on 11/17/2025).

Wei, Lai, Mei-Po Kwan, Roel Vermeulen, and Marco Helbich (Nov. 1, 2023). “Measuring Environmental Exposures in People’s Activity Space: The Need to Account for Travel Modes and Exposure Decay”. In: Journal of exposure science & environmental epidemiology 33.6, pp. 954–962. issn: 1559-0631. doi: 10.1038/s41370-023-00527-z. PMID: 36788269. url: https://pmc.ncbi.nlm.nih.gov/articles/PMC7617267/ (visited on 05/23/2026).

Wheeler, James O. (Jan. 1, 2005). “Geography”. In: Encyclopedia of Social Measurement, pp. 115–123. doi: 10.1016/B0-12-369398-5/00277-2. url: https://www.sciencedirect.com:5037/science/chapter/referencework/pii/B0123693985002772 (visited on 12/05/2025).

Zare Sakhvidi, Mohammad Javad et al. (Nov. 2025). “Methodological Guidance for Selecting Buffers in Greenspace–Health Studies”. In: The Lancet Planetary Health 9.11, p. 101370. issn: 25425196. doi: 10.1016/j.lanplh.2025.101370. url: https://linkinghub.elsevier.com/retrieve/pii/S2542519625002487 (visited on 05/10/2026).

Zheng, Lingwei, Mei-Po Kwan, Yang Liu, Dong Liu, Jianwei Huang, and Zihan Kan (2024). “How Mobility Pattern Shapes the Association between Static Green Space and Dynamic Green Space Exposure”. In: Environmental Research 258, p. 119499. issn: 0013-9351.

Zhong, Chen, Zhengzi Zhou, Nilufer Sari Aslam, Yikang Wang, and Adham Enaya (Nov. 26, 2025). “Anonymised Human Location Data in England for Urban Mobility Research”. In: Scientific Data. issn: 2052-4463. doi: 10.1038/s41597-025-06323-8. url: https://www.nature.com/articles/s41597-025-06323-8 (visited on 11/28/2025).

