## Supplementary Materials for "Accounting for Human Movement to Improve Exposure-Health Models"

**Table 1:** *Model Performance measures. Monte Carlo Error in parenthesis.*

| **Model** | **N** | **Parameter** | **Bias** | **Coverage** | **EmpSE** | **MSE** |
|---|---|---|---|---|---|---|
| Fixed-weight | 1,114 | $\beta_0$ | 0.231 (0.024) | 0.926 (0.008) | 0.768 (0.017) | 0.643 (0.028) |
| | | $\beta_1$ | -0.787 (0.065) | 0.933 (0.008) | 2.055 (0.046) | 4.840 (0.203) |
| | 50,000 | $\beta_0$ | 0.007 (0.004) | 0.956 (0.006) | 0.120 (0.003) | 0.015 (0.001) |
| | | $\beta_1$ | -0.021 (0.010) | 0.954 (0.007) | 0.318 (0.007) | 0.101 (0.004) |
| | 100,000 | $\beta_0$ | 0.006 (0.003) | 0.951 (0.007) | 0.085 (0.002) | 0.007 (0.000) |
| | | $\beta_1$ | -0.017 (0.007) | 0.944 (0.007) | 0.225 (0.005) | 0.051 (0.002) |
| Learned-weight | 1,114 | $\beta_0$ | 0.248 (0.032) | 0.859 (0.011) | 1.005 (0.022) | 1.070 (0.044) |
| | | $\beta_1$ | -0.790 (0.083) | 0.875 (0.010) | 2.612 (0.058) | 7.438 (0.331) |
| | | $\log \alpha$ | 0.043 (0.012) | 0.925 (0.008) | 0.391 (0.009) | 0.155 (0.007) |
| | 50,000 | $\beta_0$ | 0.004 (0.004) | 0.945 (0.007) | 0.142 (0.003) | 0.020 (0.001) |
| | | $\beta_1$ | -0.015 (0.011) | 0.949 (0.007) | 0.352 (0.008) | 0.124 (0.006) |
| | | $\log \alpha$ | -0.037 (0.007) | 0.931 (0.008) | 0.213 (0.005) | 0.047 (0.003) |
| | 100,000 | $\beta_0$ | 0.004 (0.003) | 0.956 (0.006) | 0.099 (0.002) | 0.010 (0.000) |
| | | $\beta_1$ | -0.011 (0.008) | 0.954 (0.007) | 0.245 (0.005) | 0.060 (0.003) |
| | | $\log \alpha$ | -0.023 (0.005) | 0.945 (0.007) | 0.149 (0.003) | 0.023 (0.001) |
| RCS | 1,114 | $\beta_0$ | 1.190 (0.025) | 0.507 (0.016) | 0.777 (0.017) | 2.019 (0.050) |
| | | $\beta_1$ | -2.948 (0.056) | 0.494 (0.016) | 1.777 (0.040) | 11.848 (0.307) |
| | | $\beta_2$ | 0.297 (0.065) | 0.932 (0.008) | 2.046 (0.046) | 4.272 (0.196) |
| | 50,000 | $\beta_0$ | 0.006 (0.005) | 0.944 (0.007) | 0.147 (0.003) | 0.021 (0.001) |
| | | $\beta_1$ | -0.019 (0.010) | 0.941 (0.007) | 0.321 (0.007) | 0.103 (0.005) |
| | | $\beta_2$ | -0.010 (0.010) | 0.954 (0.007) | 0.306 (0.007) | 0.094 (0.004) |
| | 100,000 | $\beta_0$ | 0.002 (0.003) | 0.953 (0.007) | 0.103 (0.002) | 0.011 (0.000) |
| | | $\beta_1$ | -0.007 (0.007) | 0.953 (0.007) | 0.225 (0.005) | 0.050 (0.002) |
| | | $\beta_2$ | -0.012 (0.007) | 0.960 (0.006) | 0.209 (0.005) | 0.044 (0.002) |
